# Repeatability of Pulse Oximetry Measurements in Children During Triage in Two Ugandan Hospitals

**DOI:** 10.1101/2022.12.21.22283800

**Authors:** Ahmad Asdo, Alishah Mawji, Collins Agaba, Clare Komugisha, Stefanie K Novakowski, Yashodani Pillay, Stephen Kaumu, Matthew O Wiens, Samuel Akech, Abner Tagoola, Niranjan Kissoon, J Mark Ansermino, Dustin Dunsmuir

## Abstract

**Background:** In low- and middle-income countries, health workers use pulse oximeters for intermittent spot measurements of SpO_2_. However, the accuracy and reliability when used for spot measurements has not been determined. We evaluated the repeatability of spot measurements, and the ideal observation time of measurement to guide recommendations during spot check measurements.

**Methods:** Two one-minute measurements were done for the 3,903 subjects enrolled in the study, collecting 1Hz SpO_2_ and signal quality index (SQI) data. The repeatability between the two measurements was assessed using an intraclass correlation coefficient (ICC), calculated using a median of all seconds of non-zero SpO_2_ values for each recording (any quality, Q1), and again with a quality filter only using seconds with SQI ≥ 90% (good quality, Q2). The ICC was also calculated for both these conditions using subsets of the minute, in increasing increments of 5 seconds, up to the whole minute. Lastly, the whole minute ICC was calculated with good quality (Q2), including only records where both measurements had a mean SQI > 70% (Q3).

**Findings:** The repeatability ICC with condition Q1 was 0.591 (95% confidence interval (CI) = 0.570, 0.611). Using only the first 5 seconds of each measurement reduced the repeatability to 0.200 (95% CI = 0.169, 0.230). Filtering with Q2, the whole minute ICC was 0.855 (95% CI = 0.847, 0.864). The ICC did not improve beyond the first 35 seconds. For Q3, the repeatability rose to 0.908 (95% CI = 0.901, 0.914).

**Conclusions:** Training guidelines must emphasize the importance of signal quality and duration of measurement, targeting a minimum of 35 seconds of adequate-quality, stable data. In addition, the design of new devices should incorporate user prompts and force quality checks to encourage more accurate pulse oximetry measurement.

**Trial Registration:** Clinical Trials.gov Identifier: NCT04304235, Registered 11 March 2020.

## INTRODUCTION

Pulse oximetry is a non-invasive light-through-tissue technology that uses red and infrared light to measure oxygen saturation (SpO_2_). It is commonly used in emergency and in intensive care departments, during surgery, and even at home to monitor the oxygenation status of patients. The World Health Organization’s (WHO) updated pediatric Emergency Triage, Assessment, and Treatment (ETAT) guidelines recommend using pulse oximetry to determine the presence of hypoxemia in all children with ETAT emergency signs. They also recommend oxygen supplementation if SpO_2_ levels are below 90% or 94% (depending on the presence of respiratory distress and other clinical signs).^1^ When coupled with a reliable oxygen supply, monitoring SpO_2_ with pulse oximetry in low- and middle-income countries (LMICs) has been shown to reduce mortality from pneumonia by as much as 35%.^2^ When combined with clinical signs, a low oxygen saturation is a strong predictor of the need for hospital admission,^3,4^ or death ^5^ for children with infectious illnesses in LMICs.

The durability and replacement costs of pulse oximeter sensors have been a limiting factor in sustainable implementation of pulse oximetry as a clinical tool in LMICs.^2^ The increasing recognition of the importance of supplemental oxygen therapy and the finding that the current devices often do not perform well when used by frontline health workers has catalyzed the development of new pulse oximeters better suited to low-resource settings.^6,7^ Due to limited device availability, a lack of trained personnel, and excessive patient load, spot monitoring is likely to be more acceptable than continuous SpO_2_ monitoring for children in resource-limited environments.^8^ However, the reliability of spot measurements is uncertain. One approach is to take multiple SpO_2_ readings to inform clinical decisions of high importance.^9^ However, there has been a lack of research on how to achieve the most reliable SpO_2_ value within the shortest time and with the lowest training overhead for frontline health workers.

We aimed to assess the repeatability of SpO_2_ spot measurements in routine triage of children during a clinical study in a low-resource environment, where repeatability is defined as the likelihood of getting the same results of a measurement with the same operator, same device, and same patient within a short period of time.^10^

## METHODS

### Study cohort

This is a planned secondary analysis of pulse oximetry data from an interrupted time series study to validate a digital triaging platform called Smart Triage (Clinical Trials.gov Identifier: NCT04304235).^11,12^ We obtained repeated pulse oximetry measurements from 3,903 children presenting to the outpatient department of Jinja Regional Referral Hospital (JRRH, 2,141 patients) and Gulu Regional Referral Hospital (GRRH, 1,762 patients) in Uganda. All children seeking medical treatment for an acute systemic illness during the baseline phase of Smart Triage were eligible (April 2020 to March 2021 at JRRH and March 2021 to January 2022 at GRRH). Children from birth to 19 years were recruited based on the hospitals’ age threshold for pediatric admissions. Children seeking care for elective procedures or clinical review appointments were excluded. Participation was voluntary and written informed consent was provided by a parent or guardian prior to enrollment. Assent was required from children above eight years of age.

### Data collection

Following consent and enrollment, a study nurse collected over 200 variables, including pulse oximetry and other clinical signs, symptoms, and sociodemographic variables.^13^ For each child, two one-minute pulse oximetry spot measurements were collected using a customized mobile application and a Masimo iSpO_2_® Pulse Oximeter (Masimo Corporation, USA) with micro-USB connected directly to an Android data collection tablet. The application was designed to encourage the user to obtain the highest quality recording using background color coding and forcing functions that minimized recordings of low-quality data. The user would connect the probe and wait for the color-coded quality signal, which appeared as a green background behind the waveform, before beginning the recording. Trend values, including heart rate (HR), SpO_2_, and signal quality index (SQI), were recorded at 1Hz and the raw plethysmograph waveform was recorded at 62.5Hz.^14^ The SQI was calculated as a percentage using Masimo status flags (excess light, artifacts, low perfusion, pulse search, low signal identification, and quality indicator), the perfusion index, and the variability of SpO_2_ and HR trends (Appendix A). The SQI for a spot measurement was calculated as the mean SQI from all the data. The time at the beginning of each measurement was recorded, which enabled the calculation of the time difference between the two measurements given that each measurement lasted for 1 minute. The median SpO_2_ and HR values provided by the app for each one-minute measurement were calculated using data with SQI ≥ 90% (considered good quality). If there was no continuous 30 second period of good quality data, the application prompted the user to perform an additional measurement. Any number of recordings could be done, but staff were trained to acquire two good recordings and only the two highest quality measurements were saved and then analyzed in this study.

### Data analysis

Records containing at least two pulse oximetry measurements recorded less than one hour apart and with more than 80% of their variables present were analyzed. The measurement with the highest SQI (SpO2-1) was compared to the measurement with the second highest quality SQI (SpO2-2). SpO_2_ values of 0 were ignored as these were seconds where the pulse oximeter failed to provide a value. The SQIs from the plethysmograph were summarized using histograms and the median time difference between the two recordings was calculated.

The SpO_2_ for the spot measurements was re-calculated from the 1Hz data in three different ways, referred to as Q1-Q3. First, SpO_2_ dataset Q1 was calculated from the 1Hz data using all records and all non-zero seconds of SpO_2_ data. This calculation is most reflective of real-world clinical applications in which devices provide SpO_2_ values whenever they can be calculated and frontline health workers use numbers directly from the devices, without any intermediate averaging. The second SpO_2_ dataset, Q2, included all records, and followed the data collection application rules of only using data with good quality (an SQI ≥ 90%). This calculation indicates the repeatability under optimal SQI conditions while not excluding any patients. The third SpO_2_ dataset, Q3 applied the good quality data rule from Q2, but only included records that had a mean SQI value (from all the data) of above 70% for both SpO2-1 and SpO2-2.

Agreement between the two SpO_2_ measurements using dataset Q1 were visualized using Bland Altman plots (SpO_2_ difference vs mean of the two measurements) to identify any systematic error within the measurements and possible outliers. Outliers were visually identified from the plots and were defined as measurements with an SpO_2_ absolute difference of greater than 20%. The repeatability bias (see Table 1 for definitions) and the limits of agreement were calculated for datasets Q1 and Q2.

**Table 1.** Definitions of the terms used in the paper derived from the International Organization of Standardization ^15^.

| Term | Definition |
| --- | --- |
| Precision | The closeness of agreement between independent test results obtained under stipulated conditions. |
| Repeatability | Precision under repeatability conditions. In this study, this is the repeatability of pulse oximetry (SpO<sub>2</sub>) in children. |
| Repeatability conditions | Conditions where independent test results are obtained with the same method on identical test items in the same laboratory by the same operator using the same equipment within short intervals of time. |
| Repeatability bias | The bias under repeatability conditions, considering what set of samples as the test and the other as the reference. In this study, for the purposes of bias, the highest quality SpO<sub>2</sub> recordings (SpO2-1) are considered the reference value and the second highest quality recordings (SpO2-2) are considered the test results. |
| Accuracy | The closeness of agreement between a test result and the accepted reference value. |
| Bias | The difference between the expectation of the average test results and an accepted average reference value. |

Repeatability (see Table 1) was assessed using the intraclass correlation coefficient (ICC) between the median SpO_2_ from SpO2-1 and SpO2-2 using a two-way random effects model with a single rater. The ICC was calculated for all three datasets, and the (Q1) and (Q2) calculations were repeated for different spot measurement durations in increments of 5 seconds up to the full minute of recording. Some patients for some durations had zero seconds of usable (non-zero SpO_2_) data and thus were not included in the ICC calculation. We then plotted these two conditions as overlapping bar graphs to see the effect of signal quality and measurement duration on the repeatability. Data was analyzed using R software version 4.1.2.^16^

### Ethics considerations

This study was approved by the institutional review boards at the University of British Columbia in Canada (ID: H19-02398; H20-00484), the Makerere University School of Public Health in Uganda and the Uganda National Council for Science and Technology.

## FINDINGS

Of the 3,903 patients enrolled in the study, we excluded 80 patients due to missing paired measurements (58) or time difference between the measurements of longer than one hour (22), leaving 3,823 patients who were included in this analysis. The median age (Interquartile range (IQR)) was 15 (26.6) months and 52% were male (Table 2).

**Table 2.**
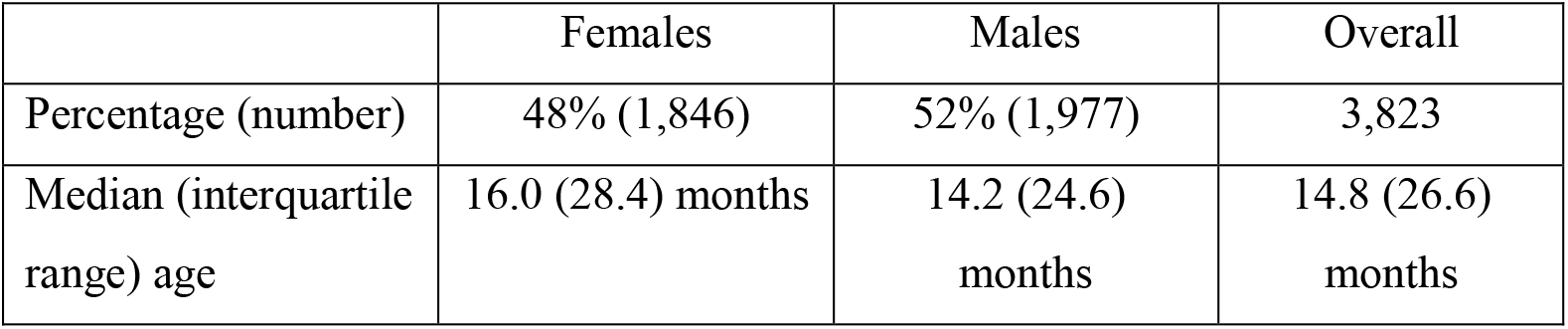
Demographic variables of the patients included in the study.

|  | Females | Males | Overall |
| --- | --- | --- | --- |
| Percentage (number) | 48% (1,846) | 52% (1,977) | 3,823 |
| Median (interquartile range) age | 16.0 (28.4) months | 14.2 (24.6) months | 14.8 (26.6) months |

The median (IQR) SQI for all the data was 93% (92-97%) (Fig 1). Most (93%) of the highest-quality observations (SpO2-1) from the two paired spot measurements had SQI ≥ 90%. For the lower quality observation (SpO2-2), 78% of measurements had SQI ≥ 90. There was a median (IQR) of 0.21 (0.13 - 0.65) minutes between measurements and 95% of the measurements occurred less than 8 minutes apart.

**Figure 1.**
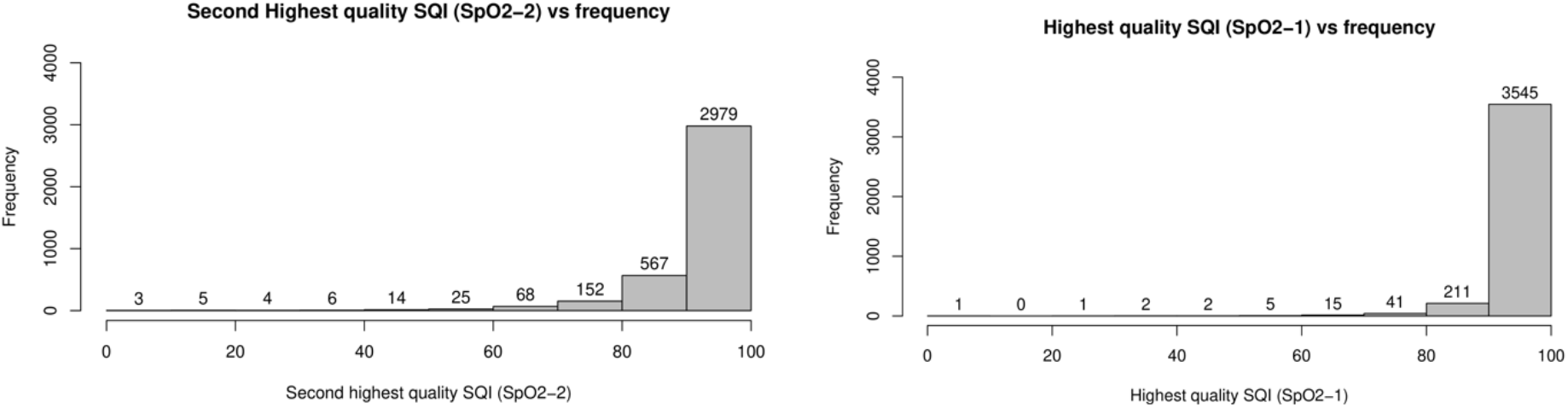
Histogram of oxygen saturation measurements for SpO2-1 and SpO2-2.

In dataset Q1 (no quality consideration), both SpO2-1 and SpO2-2 had a median SpO_2_ ≥ 90% for 99.0% of the records. Two patients were identified as outliers and both these patients had hypoxemia (SpO_2_ < 90%) for one of the two measurements.

Large disagreements between SpO2-1 and SpO2-2 were only seen when the mean SpO_2_ of the two measurements was low (Figure 2). The repeatability bias of SpO_2_ was 0.04%. The spread between the upper and lower limits of agreement was 5.8%. Using Q2, the repeatability bias was 0.03% and the spread of the upper and lower limits of agreement was 5.2%.

**Figure 2:**
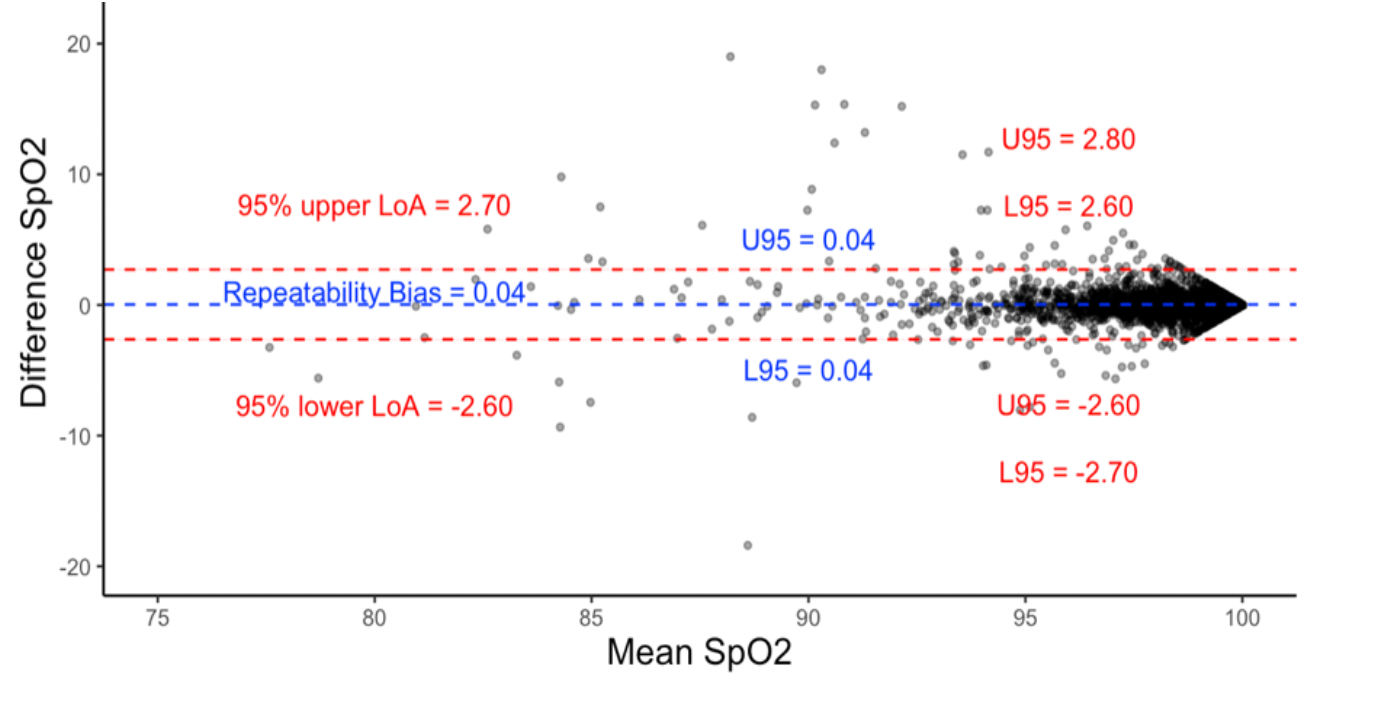
Bland Altman plot for SpO_2_ values. The total number of cases (N) = 3,821. Each data point is for a pair of recordings with the y-axis as difference (SpO2-1 – SpO2-2) and the x-axis as the mean SpO_2_. Two outliers excluded. Incudes only SpO_2_ > 0. U95 = upper limit of confidence interval, L95 = lower limit of confidence interval. LoA = Limit of agreement.

The ICC (Q1) (95% CI) for SpO_2_ measurements for the 60 seconds duration was 0.58 (0.56, 0.6) (Figure 3). The ICC (Q1) for the median (95% CI) SpO_2_ for only the first 5-seconds was 0.2 (0.17, 0.23). In general, the ICC (Q1) value increased with increasing duration of up to 40 seconds. After 40 seconds, the ICC (Q1) plateaued at 0.5. The ICC (Q2) was much larger than the ICC (Q1) for all durations except in the first 10 seconds when the repeatability was very low for both. The ICC (Q2) plateaued after 35 seconds at 0.85. For the full 60 second duration, the ICC (Q3) (95% CI) was even higher at 0.91 (0.9, 0.91); (n= 3,230).

**Figure 3:**
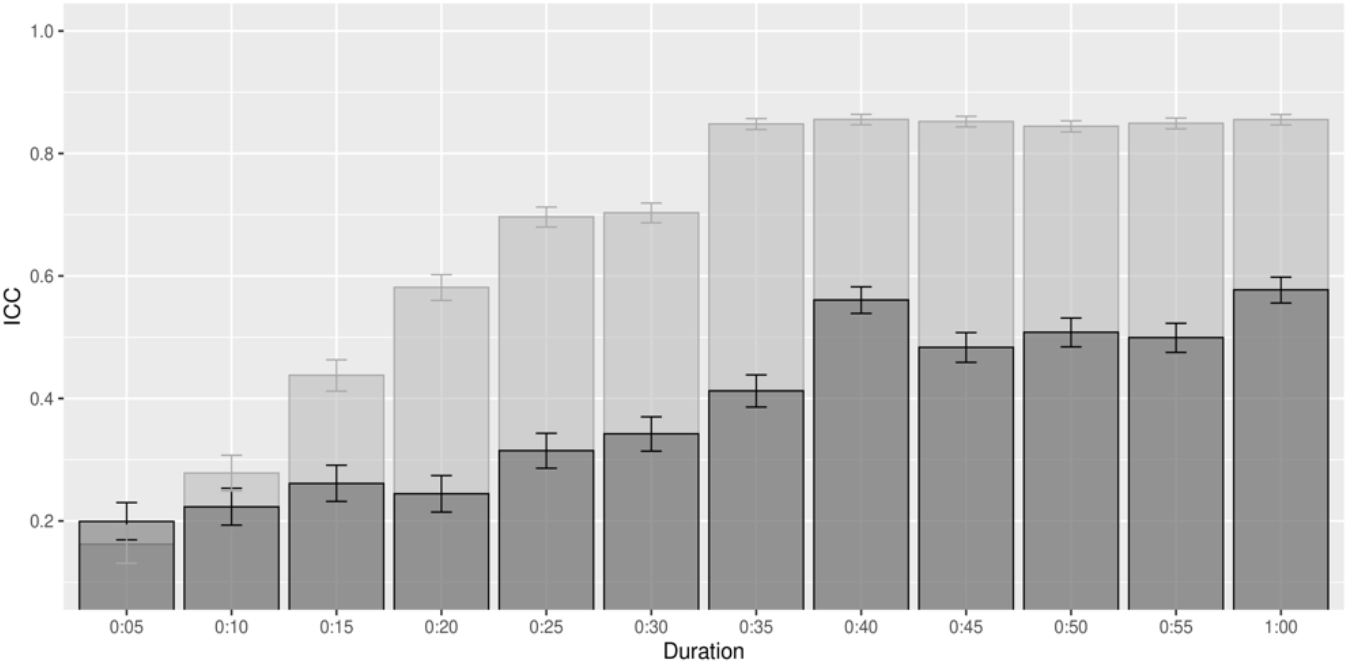
ICC (Q1 and Q2) vs Duration. The total number of cases (N) = 3,823. Dark bars represent intraclass correlation coefficient (ICC) when including all seconds regardless of their SQI value (Q1). Light bars represent ICC when only seconds with SQI ≥ 90% were included (Q2). Less than 1% of the records were excluded in the first 15 seconds since those records did not have enough seconds matching the quality criteria. The bars indicate the ICC values over varying spot measurement durations.

## DISCUSSION

### Summary

In this manuscript we report the repeatability of pulse oximetry measurement in children during triage at two hospitals in Uganda based on two 60-second spot measurements of SpO_2_ per child. The repeatability value for the full minute (ICC 0.85) was representative of high repeatability; however, shorter periods of observation, lower signal quality, and lower oxygen saturation reduced repeatability. When low quality data was included, there was a large reduction in repeatability and measurement agreement. Repeatability increased as the duration of recordings lengthened until the ICC plateaued at 35 seconds. Improved repeatability of SpO_2_ recordings can be achieved with practices such as optimizing signal quality, extending the duration of the recording to at least 35 seconds and by performing repeat observations when SpO_2_ measurements are below 90%.

### Implications for clinical care

The expansion of digital health tools in LMICs has been accelerated by the COVID-19 pandemic.^18,19,20^ Pulse oximeters are in demand in low resource settings, and it is critical that they are used appropriately.^8,21^ The integration of pulse oximetry into the digital health landscape is an opportunity to design intelligent systems that include averaging for spot measurements, filtering based on quality criteria, and enforcing of minimum measurement durations. Understanding the factors that reduce repeatability will facilitate improved device design and optimize clinical procedures and training.

We have shown that both SQI and duration of measurement should be optimized to improve repeatability. We observed that without quality filtering, two SpO_2_ measurements done by the same observer, using the same device on the same patient over a short period of time, have only moderate repeatability based on an ICC of 0.5. There are multiple factors that will reduce the repeatability of an SpO_2_ measurement in addition to the accuracy of the device itself. These factors include the short-term variation within the patient (within subject variation), the variations in how observers perform the measurement (within and between observer variation), and other variations in filtering, averaging, or rounding of measurements.

It may take time for the measurement to stabilize when applying a pulse oximeter to a child due to the delay in signal processing algorithms in the device.^22^ This period of stabilization will vary between devices, subjects (based on their oxygenation status), and operators. Poor application of the sensor (sensor exposure to external light) or a restless child (sensor motion) are both recognized by the Masimo pulse oximeter and contributed to lower SQI and thus delays in stabilization of the measurement. However, we have shown that choosing an observation obtained over a short time frame (initial 5 seconds) will lead to poor repeatability and larger variability than a longer measurement, even when data collectors are trained to wait for this stabilization and design affordance features are used (i.e., a green colored background indicating good quality data, SQI ≥ 90%). The increased variability will most likely lead to reduced accuracy in the measurement, which can ultimately affect prompt identification and treatment of hypoxemia.

### Limitations

This study was performed at only two institutions and included only 14 observers. All pulse oximeter data was collected with identical pulse oximeter models and a custom application that was designed to improve observer performance. There may have been some dependency between observers, but initial analysis of the ICC (Q1) using a multiple-rater model showed no significant difference, so this dependency was not included in the final analysis. Further, external validity of the study may be limited as the study sites in Uganda are at an altitude of 1100-1200m above sea level. The relative hypoxia at this altitude may have reduced the repeatability due to the non-linear shape of the oxygen saturation curve. The range of SpO_2_ values measured may have biased ICC values compared to observations at sea level. The largest differences in SpO_2_ were seen at low SpO_2_ values, but this is to be expected given that the majority of the SpO_2_ values were high (>90%) and on the flat part of the oxygen dissociation.^17^ Additionally, this secondary analysis did not consider clinical outcomes, nor attempt to define an acceptable ICC value for adequate repeatability. ICC values have previously been defined as the following: less than 0.2 slight repeatability, between 0.2 and 0.4 low repeatability, between 0.4 and 0.7 moderate repeatability, between 0.7 and 0.9 high repeatability, and greater than 0.9 very high repeatability. ^23^ Using this scale, this study showed that repeatability increased from low to moderate when using 35 seconds of data versus a shorter time frame. A further increase to high repeatability occurred when only good quality data was used to calculate the median reported SpO_2_. Lastly, poor repeatability of pulse oximetry measurements may not necessarily indicate poor accuracy, and one should not be taken as a substitute for the other.

## Conclusion

This study demonstrates that shorter periods of observation, lower signal quality, and lower oxygen saturation levels reduce repeatability of pulse oximetry measurements. Repeatability of observations is critically important for making optimal clinical decisions but also essential when performing device validation. These results should inform training for health workers who perform pulse oximetry. For example, observers (or a pulse oximetry device) should average SpO_2_ readings for at least 35 seconds of good quality data to obtain a repeatable measurement. Future studies should concentrate on within observer uncertainty in different settings and with different devices and evaluate other causes of uncertainty such as within subject variability.

## Data Availability

All data analyzed in the study are available upon reasonable request to the authors

## Acknowledgements

We would like to thank the Jinja and Gulu Regional Referral Hospital Smart Triage research staff for collecting the data, all children and caregivers who took part in the study, and biostatistician Cherri Zhang for statistical advice.

## APPENDIX A

The Signal Quality Index (SQI) calculated during the pulse oximetry data collection for the Smart Triage study has a range of 0-100%. 100% is considered to be a perfect quality signal. The algorithm takes the following input at 1Hz: HR (heart rate), SpO_2_ (oxygen saturation), PFI (perfusion index) and Masimo flags for no sensor detected, defective sensor, sensor off patient, search for pulse, low Signal IQ, and low perfusion.

The SQI algorithm starts with 100% and subtracts penalty values from this based on any conditions identified. The SQI is never set below 0%.

Pseudo code for the algorithm (run at 1Hz):

1. If there is no HR calculated (HR = 0), `SQI = 0`, skip all below steps.
2. Set `SQI = 100`.
3. Check the Masimo flags:
  a. If no sensor detected, defective sensor, or sensor off patient, `SQI = 0`.
  b. If artifacts flag within the last 1.2 seconds, `SQI = SQI – 40`.
  c. If searching for pulse, `SQI = SQI – 60`.
  d. If low perfusion flag, `SQI = SQI – 50`.
4. Calculate PFI as a percentage out of 20.
  a. If `PFI% ≤ 2.5%` then `SQI = 0`.
  b. Otherwise, if `PFI%` ≤ `6%` then `SQI = SQI – 40`.
  c. Otherwise, if PFI% < 25% then give a small penalty according to the following: Calculate P as the percent of the way from 25% down to 6% using the formula: 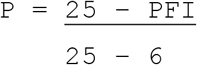 Then multiply this value by 10 to get PP (0 < PP < 10), then `SQI = SQI – PP`.
5. Check SpO_2_ variability. The last 30 seconds of SpO_2_ data is stored to look at variability.
  a. Calculate the interquartile range of SpO_2_, `SIQR`.
  b. Calculate the median SpO_2_, `SM`.
  c. If 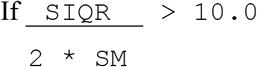, then `SQI = SQI – 40`.
6. Check HR variability. The last 30 seconds of HR data is stored to estimate variability.
  a. Calculate the interquartile range of HR, `HIQR`.
  b. Calculate the median HR, `HM`.
  c. If 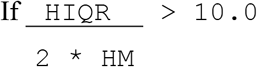, then `SQI = SQI – 40`.

The resulting SQI from the above algorithm is displayed as a color in the data collection app. Below is a diagram of the colors used. Only green seconds of data (SQI ≥ 90) are used in calculating an overall median HR and SpO_2_ for the spot-check measurement (Figure 4).

**Figure 4:**
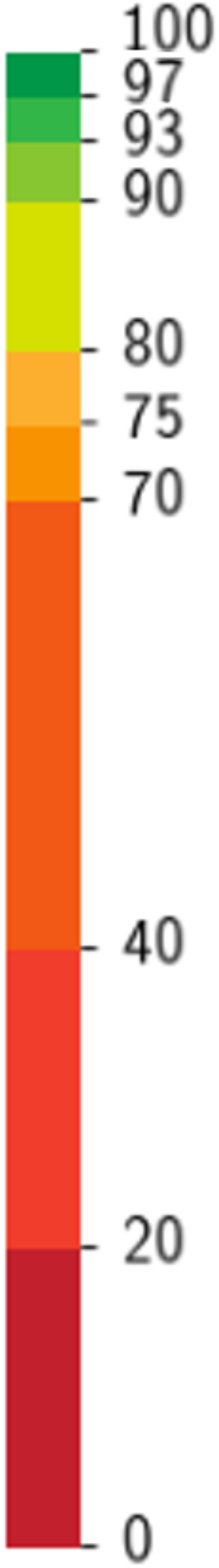
SQI color coding feedback in the data collection app.

## Bibliography

1. World Health Organization. Paediatric Emergency Triage, Assessment and Treatment UPDATED GUIDELINE.; 2016. Accessed November 22, 2022. https://apps.who.int/iris/handle/10665/204463

2. Duke T, Graham SM, Cherian MN, et al. Oxygen is an essential medicine: a call for international action. Int J Tuberc Lung Dis. 2010;14(11):1362–1368.

3. Raihana S, Dunsmuir D, Huda T, et al. Correction: Development and internal validation of a predictive model including pulse oximetry for hospitalization of Under-five children in Bangladesh. PLOS ONE. 2016;11(1). doi:10.1371/journal.pone.0147560

4. Mawji A, Akech S, Mwaniki P, et al. Derivation and internal validation of a data-driven prediction model to guide frontline health workers in triaging children under-five in Nairobi, Kenya. Wellcome Open Research. 2021;4:121. doi:10.12688/wellcomeopenres.15387.3

5. Reed C, Madhi SA, Klugman KP, et al. Development of the respiratory index of severity in children (RISC) score among young children with respiratory infections in South Africa. PLoS ONE. 2012;7(1). doi:10.1371/journal.pone.0027793

6. Herbert LJ, Wilson IH. Pulse oximetry in low-resource settings. Breathe. 2012;9(2):90–98. doi:10.1183/20734735.038612

7. Baker K, Akasiima M, Wharton-Smith A, et al. Performance, acceptability, and usability of respiratory rate timers and pulse oximeters when used by frontline health workers to detect symptoms of pneumonia in Sub-Saharan africa and Southeast Asia: Protocol for a two-phase, multisite, mixed-methods trial. JMIR Research Protocols. 2018;7(10). doi:10.2196/10191

8. Duke T, Subhi R, Peel D, Frey B. Pulse Oximetry: Technology to reduce child mortality in developing countries. Annals of Tropical Paediatrics. 2009;29(3):165–175. doi:10.1179/027249309x12467994190011

9. Karlen W, Petersen CL, Dumont GA, Ansermino JM. Variability in estimating shunt from single pulse oximetry measurements. Physiological Measurement. 2015;36(5):967–981. doi:10.1088/0967-3334/36/5/967

10. Harper DGC. Some comments on the repeatability of measurements. Ringing & Migration. 1994;15(2):84–90. doi:10.1080/03078698.1994.9674078

11. Mawji A, Li E, Komugisha C, et al. Smart triage: Triage and management of sepsis in children using the point-of-care pediatric rapid sepsis trigger (PRST) tool. BMC Health Services Research. 2020;20(1). doi:10.1186/s12913-020-05344-w

12. Mawji A, Li E, Dunsmuir D, et al. Smart triage: Development of a rapid pediatric triage algorithm for use in low-and-middle income countries. Frontiers in Pediatrics. 2022;10. doi:10.3389/fped.2022.976870

13. Mawji A, Li E, Chandna A, et al. Common data elements for predictors of pediatric sepsis: A Framework to standardize data collection. PLOS ONE. 2021;16(6). doi:10.1371/journal.pone.0253051

14. Dunsmuir D, Petersen C, Karen W, Lim J, Dumont G, Ansermino JM. THE PHONE OXIMETER FOR MOBILE SPOT-CHECK. 2012.

15. ISO 5725-1:1994. ISO. https://www.iso.org/standard/11833.html. Published April 14, 2022. Accessed November 23, 2022.

16. The R project for statistical computing. R. https://www.r-project.org/. Accessed November 23, 2022.

17. Jubran A. Pulse oximetry. Critical Care. 2015;19(1). doi:10.1186/s13054-015-0984-8

18. Mitgang EA, Blaya JA, Chopra M. Digital Health in response to COVID-19 in low- and middle-income countries: Opportunities and challenges. Global Policy. 2021;12(S6):107–109. doi:10.1111/1758-5899.12880

19. Silenou BC, Nyirenda JL, Zaghloul A, et al. Availability and suitability of digital health tools in Africa for pandemic control: Scoping review and cluster analysis. JMIR Public Health and Surveillance. 2021;7(12). doi:10.2196/30106

20. Budd J, Miller BS, Manning EM, et al. Digital Technologies in the public-health response to COVID-19. Nature Medicine. 2020;26(8):1183–1192. doi:10.1038/s41591-020-1011-4

21. Tuti T, Aluvaala J, Akech S, Agweyu A, Irimu G, English M. Pulse oximetry adoption and oxygen orders at paediatric admission over 7 years in Kenya: A multihospital retrospective cohort study. BMJ Open. 2021;11(9). doi:10.1136/bmjopen-2021-050995

22. Ward R, Raheja P, Singh A. Comparison of Latencies and Time to Stabilization of Pulse Oximeters at a Tertiary Health Care Facility. Int J Sci Stud 2015;3(6):170–174

23. Hassan AT, Ahmed SM, AbdelHaffeez AS, Mohamed SAA. Accuracy and precision of pulse oximeter at different sensor locations in patients with heart failure. Multidisciplinary Respiratory Medicine. 2021;16. doi:10.4081/mrm.2021.742

